# Early guidance for Sars-Cov-2 health policies in India: Social Distancing amidst Vaccination and Virus Variants^*^

**DOI:** 10.1101/2022.02.02.22270353

**Authors:** Yi Zhang, Sanjiv Kapoor

**Author notes:** Corresponding author,. Part of this work was performed when S. Kapoor was a visiting Professor at IIT-Delhi. Research sponsored in part by NSF. grant No. 2028274.

## Abstract

Policy decisions during the SARS-COV-2 pandemic were complicated due to virus variants and the impacts of societal restrictions. Accurate predictive models were required in this context. In this paper we report results from a model that helped in predicting the impact of SARS-CoV-2 virus transmission in India over a period of a number of months from June, 2021 to March 2022. These models were applied in the context of enabling policy decisions to tackle the impact of the pandemic in India culminating in early warning projections for the Omicron variant and used for advise on preemptive policy actions.

**Methods:** Our model utilizes a deterministic compartment models incorporating a dynamic transmission factor, dependent on the population’s behavior as a function of the reported confirmed cases of virus transmission as well as methods for estimation of the increase in susceptible population when social distancing mandates are relaxed. The model used to predict viral growth incorporates the state of vaccination and the virus variants that form part of the transmission dynamics as well the lockdown state of the population. NPI actions were used in India to contain the spread of infections during the period of study, especially during the surge of the Omicron variant of the virus. Further we present the impact of lockdown policies and illustrate the advantage of adopting partial lockdown policies in the early period of 2022. Based on the models, our predictive analysis, when applied to the Omicron variant, illustrated substantial improvement even when partial lockdown is planned.

**Findings:** This report presents models and results that incorporated the impact of vaccination rates and the Omicron variant and were used to establish projections on the growth of Sars-Cov2 infections in India for the period from July 2021 till March 2022. The growth rate of the Omicron virus was deduced from data that originated from South Africa in November 2021. These projections were submitted to a pivotal government organization involved in developing a national public health strategy to address the pandemic and, as per personal communication, were considered when formulating national policy. The pandemic had a subdued impact in India during the period from July 2021 till date as evident from the deaths reported by the government. The projections were made every month and cases were projected over the next 4-16 weeks. The projections of cumulative cases during the Omicron wave had low errors when measured using RMSE per capita and had a MAPE error of 17.8% when measured 15 days after start of the projection on December 5th, 2021.

**Discussion:** The composed model was found to be useful in providing predictive and data based analytic input to inform early warning approaches in the context of policy based interventions to control the pandemic in India. The model provided monthly early prediction of the spread and impact of the SARS-COV-2 virus in India, state-wise, during the phase of removal of government lockdown in the second half of 2021. The early warning system incorporated the impact of the Omicron variant to provide predictions for Indian states and the country.

## 1 Introduction

The SARS-COV-2 virus has been plaguing the world since January 2020, compounding the original problem with multiple variants. Containment measures like social distancing are key, even in the presence of vaccinations, to preventing the spread and also to contain variants. Accuracy in models that predict the growth in infections are very important. While short term analysis of a few weeks is typically more accurate, long range models are important in planning. Our motivation was the impact of the virus in India during March to May 2021, in this paper we consider a model for virus infection spread that accounts for the impact of lockdown policies, vaccinations and the impact of vaccine-resistant virus variants.

Effective public health policy requires timely social distancing guidelines; as such it is critical to develop early-warning systems that would predict the rise of the virus, especially as lockdown or social distancing policies are relaxed. These warning systems would include parameters that reflect the relaxation of NPI practices related to masking, enhanced mobility etc. that would be indicative of behavior patterns that create an influx of susceptible into the population. Since the incubation period of the disease and the spread in asymptomatic population, especially the vaccinated population, results in delayed impact on the progression of the disease, long term modeling of the spread of the virus is critical.

This report deals with an analysis of the spread of infection as more population emerges from lockdown and adopts lax behavior with respect to social distancing. Impact of relaxed norms are apparent in the multiple waves that have afflicted countries around the world. We construct a model that reflects the vaccination status of the population, accounting for the time-dependent efficacy of the vaccines, and also incorporates virus variants. The analysis is based on epidemic spread models that utilize compartment models which have along history (see [6, 9] and [10, 11, 12]). A key differentiation in our work is the use of dynamic social distancing factor in the model, termed the SIR-SD model introduced in [25]. The role of reducing contacts to reduce transmission is well studied [24, 7, 13] as more recent modeling efforts have focused also on mobility [5], amongst other factors [15, 21, 14, 22, 3, 18, 19, 16, 23]. Our study incorporates virus variants. The compartment models we utilize includes compartments reflecting the various states of the population including vaccinated population, population under lockdown etc. As mentioned above, we also use a social distancing factor to modify the virus transmission rate that reflects the behavior of the population as the number of infection cases rises and reporting of the infections, as well as government mandates, becomes more prevalent. As introduced earlier in [25] the transmission of the virus incorporates a factor that is inversely dependent on the number of cumulative infections incorporated by a time-dependent susceptible population size.

This model had been utilized for multiple months, starting from June 2021, in order to provide predictive scenarios months in advance. We calibrated resurgence of the virus in our model using a release of population into the susceptible pool during the second wave of infections in India in April 2021. The wave had come as a surprise to policy makers and we designed a predictive long range model since it was important to ensure that the infection growth was contained as lockdown was removed and vaccination progressed. Our model incorporates a population that transfers from a lockdown state to a susceptible state. To quantify the growth of the susceptible population in our model, we assessed the growth of susceptible population, *H*, that led to the second wave and based future projections on this baseline behavior estimate, using this growth to determine the rate at which the population entered the susceptible set, as a result of relaxed government policies and social distancing behavior. We report modeling two rates of relaxation of lockdown procedures after the second wave that direct the rate of policy changes after June 2021: a fast rate of release where the susceptible population *H* was released in 30 days; a moderate or medium rate of release where the susceptible population *H* was released in 45 days. We illustrate the succession of projections for the fast release rate (30 day release) in Figure 1. The projections range is bounded by two scenarios, an optimistic and a pessimistic scenario. The two scenarios differ in the estimates of the addition to the susceptible population resulting from the relaxation of the lockdown that was initiated in June 2021. Over the period we studied, projections showed a reduction in cases and then an increase during the winter months ending in a sharp rise in predicted cases when Omicron started and became prevalent. Our initial lower projections of the growth of infections during August, September and October could possibly be attributed to government actions including slow release from lockdown state and enhanced vaccination rates. Our projections in December, 2021 appear higher than the actual confirmed cases as likely the rise in cases was tempered by government policies, that included night curfews as well as business and school closings during January. The dynamic progression of infection case projections is illustrated in the accompanying [video](https://drive.google.com/file/d/1ohzgOveRDM5Ncgh85mN42Metsxee1Zbo/view)[2]. Detailed estimates provided in later sections illustrate the mixed impact of the two variants, with the Omicron variant rapidly dominating the currently existing Delta variant. In later sections in this paper we also consider the impact of lockdown on reduction in viral spread.

**Figure 1:**
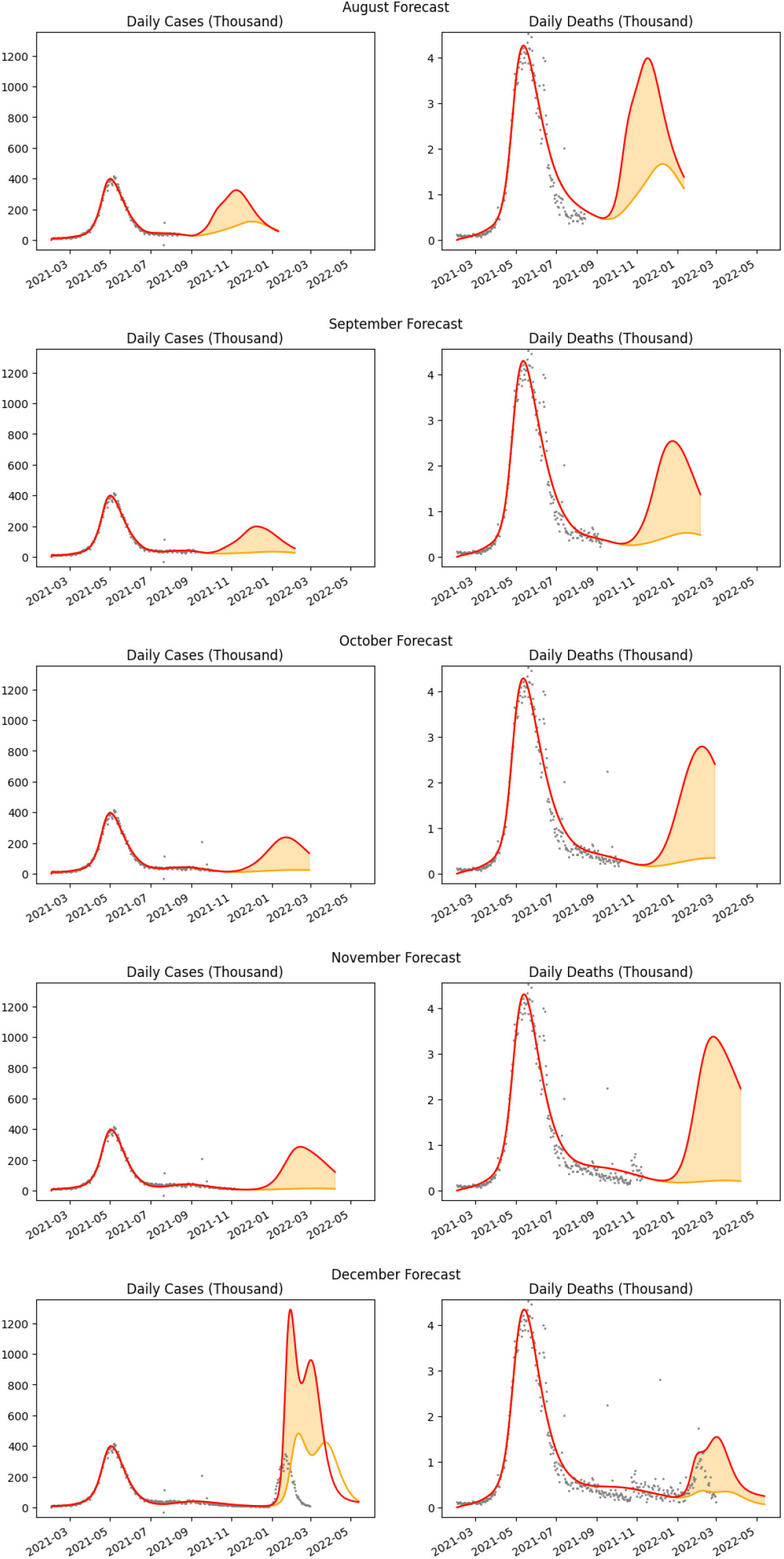
Monthly sequence of projections for the case of fast release rate. The peak case load varied according to the current infection trends and successively delayed by almost a month.

This work was motivated by the problem of determining policy decisions as vaccinations progressed and we used this model to provide monthly updates on projections to a pivotal government organization. The projections focused on the onset of the third wave and covered the period from June 2021 till December 2021. The third wave in India started in December 2021. As per personal communication these projections were considered when formulating national policy. The policies implemented at state levels included mask mandates, physical school closings, partial curfews and office closings, an example provided by Delhi state government policies [1]. The projections were accompanied by data on social distancing and mask usage from IHME. However we did not use these social distance metrics as part of our model as this data was typically delayed.

Accuracy of our model was established during the initial part of study, from April to May 2021. The changing governmental policies on non-pharmaceutical interventions, as and when rise in infection cases were evident, made accuracy analysis for the projections difficult. However, during the Omicron variant wave the confirmed cases of viral infection were substantial and an accuracy analysis of our projections versus the actual confirmed cases, showed that the cumulative case counts had very low RMSE per capita (0.0013 per capita, and had MAPE errors ranging from 17.8% to 41.6% for projections 15 to 45 days from December 15, 2021. As time progressed the deviation from our projections grew due to the NPI policies that were implemented.

While the cases incidences were kept limited till November, relaxed social distancing norms and waning vaccine efficacy likely had an impact on the cases incidences in December 2021, along with increased transmissibility of the Omicron variant.

## 2 SEIAR-SD-L Model

We use a mechanistic model as illustrated in Figure 2. Apart from standard compartments representing the Susceptible *S*, Exposed *E*, Infected (confirmed via testing) *I*, Undetected Symptomatic and Asymptomatic *A*, we include hospitalized *I*_*H*_ and home treated *I*_*N*_, along with dead *D* and recovered *R*. There was no data on deaths due to undetected symptomatic cases and so we omit the corresponding transition.

**Figure 2:**
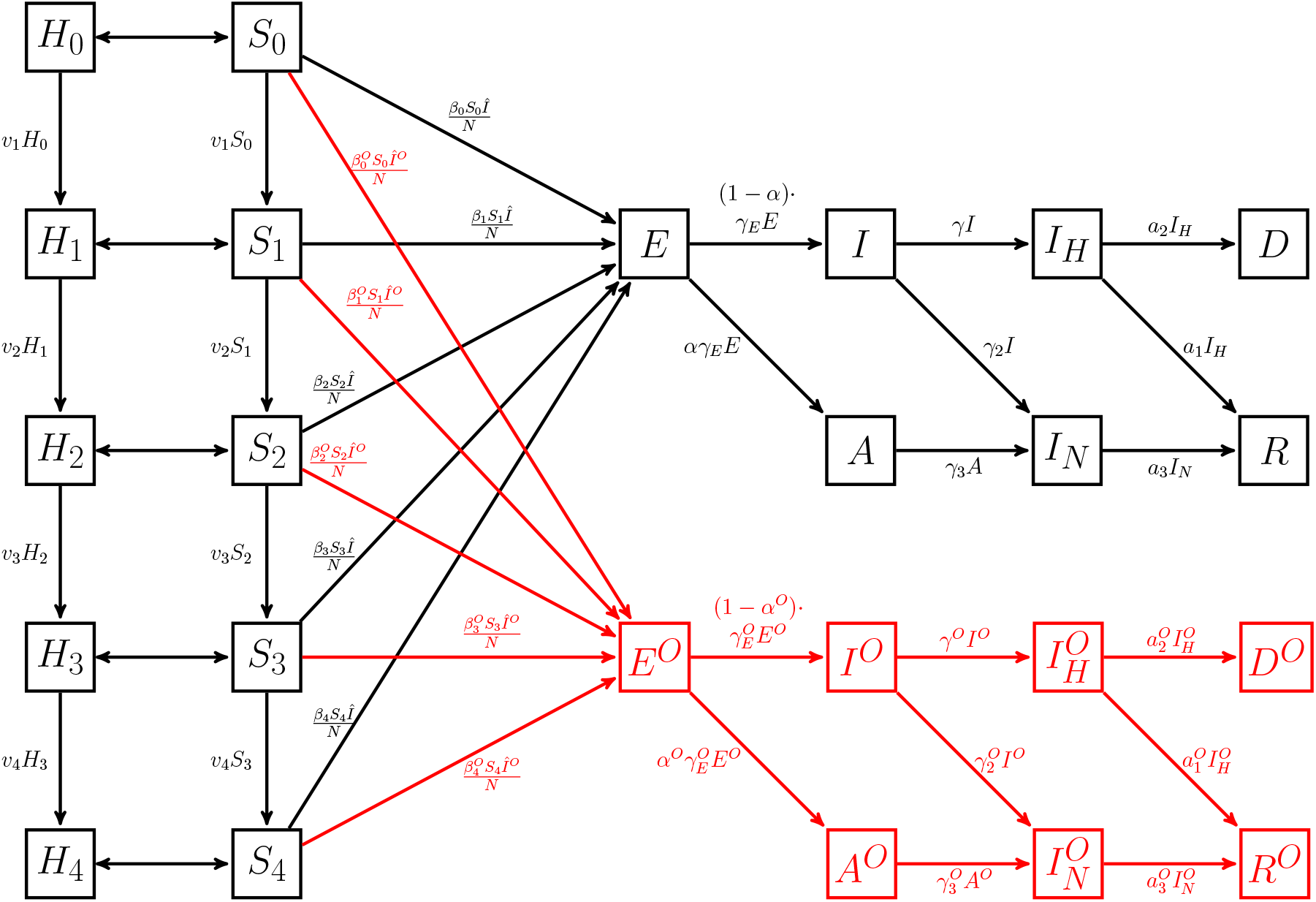
The SEIAR-SD-L Compartment Model with vaccination and two strains of the virus; delta and omicron (red)

Each of these compartments are replicated for various types of virus variants to model the impact of each type of virus variant. In the context of the Omicron virus in addition to the base variant, the Delta variant, these compartments are indexed by superscript *O*. Additionally we have a population that is on lockdown, grouped by different vaccination states, *H*_0_: the set of unvaccinated population under lockdown; *H*_1_: the population that has received the first shot of vaccination (based on the rate of vaccination of the population); *H*_2_: population after two weeks of receiving the first dose(first dose effective two weeks after the dose) ; *H*_3_: population after two weeks of receiving the second dose of vaccination given after a fixed delay from the first dose); *H*_4_: vaccinated population after 4 months of the second dose. Vaccination efficacy has been known to drop by 50% after 4 months and *H*_4_ models the population in this state [4]. Similarly the vaccination status of the susceptible population is modeled by *S*_0_, *S*_1_, *S*_2_, *S*_3_ and *S*_4_.

A release from lockdown transfers population from the corresponding lockdown compartments to the susceptible population, while lockdown policies transfer susceptible back to the *H* compartments.

### 2.1 The Compartment Model

We let

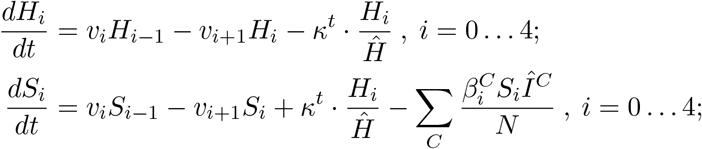

where *Ĥ* = *H*_0_ + *H*_1_ + *H*_2_ + *H*_3_ + *H*_4_, *v*_0_ = 0, *v*_5_ = 0, and *v*_1_, *v*_2_, *v*_3_, *v*_4_ represent transition rates between the population groups as represented in Figure 2. And for all strains, *C*, the following conditions hold, where we omit the notation for the *C* for ease of understanding.

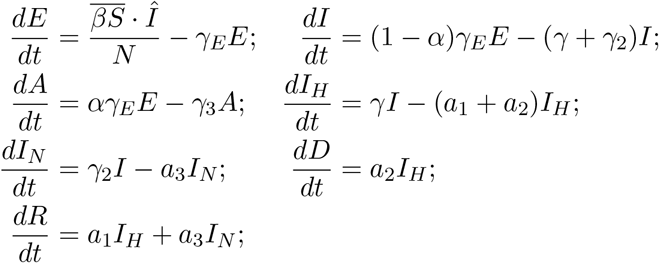

where 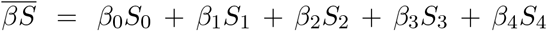 and *Î* = *I* + *A*. The model parameters, *γ*_*E*_, *γ, γ*_2_, *α, a*_1_, *a*_2_, *a*_3_ are all with respect to the strain *C*.

To model the impact of the virus on the population’s behavior we utilize dynamic transmission factor *β* [25]. The value of *β* is based on the following generic transmission function that models the dynamics of population behavior (for further discussion see [25]):

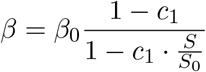

where *S* is the number of the current susceptible population and *S*_0_ the original set of susceptible population.

The function *κ*^*t*^ varies dependent on whether the policies are in a release phase or in a lockdown phase. We use a linear function in our model:

- Release Phase: In the release phase, between time *t*_1_ and *t*_2_, the function *κ*^*t*^ = *h*^*R*^, *t*_1_ ≤ *t* ≤ *t*_2_ where *h*^*R*^ is a positive constant and where *t*_1_ and *t*_2_ is the range of time over which the population enters into a susceptible state. The value of *h*^*R*^ is determined by the release policies and adherence.
- Lockdown Phase: In the lockdown phase, between time *t*_3_ and *t*_4_, the function *κ*^*t*^ = *h*^*L*^, *t*_3_ ≤ *t* ≤ *t*_4_ where *h*^*L*^ is a negative constant, represents the rate at which the population withdraws into a state of lockdown, over the range of time [*t*_3_, *t*_4_]. The value of *h*^*L*^ is determined by the strength of the lockdown and adherence.

The theoretical value of *R*_0_ in this model is analyzed by the method provided in the supplementary material and is:

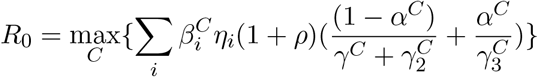

where *ρ* = *κ*^*t*^/∑ _*i*_ *S*_*i*_ is the rate at which population enters the susceptible set *S*_*i*_, *η*_*i*_ is the fraction of population in susceptible set *S*_*i*_.

## 3 Model based Projections

The SEIAR-SD-L model was used to create projections in order to help guide policy advice.

While successive projections illustrated in Figure 1 had been communicated periodically to Niti Ayog, we provide details here of the projections that include both the delta and the highly transmissible Omicron variant. Given that substantial time had elapsed since the start of the vaccination program, we utilized vaccination rate efficacy to include breakthrough cases.

These projections on the Sars-Cov2 crisis were based on infection case data until December 15th, 2021. We estimated the transmissibility of Omicron variant from the South African data of the virus and comparing the transmissible factor with the previous variant to compute a beta factor that was used to determine the transmission factor that is used for Omicron. The fitting for South Africa is detailed in the supplementary material (Figure S3). In the absence of data on incubation period and infectious period we utilized the parameters of the delta variant as estimated from our model during the phase where the delta variant was prevalent. The vaccine efficacy is adjusted for and is considered less effective w.r.t. Omicron. We present results that are parameterized on the vaccine efficiency and listed below.

### Baseline Behavior and Assumptions

We established projections that were based on the initial growth in population that were part of transmission dynamics in the first half of 2021 between February and June (first phase), and utilize that to create a baseline for the release of population from lockdown during February-March 2021 when restrictions were removed in late January. This estimate of the release with *κ*^*t*^ = *h*^*L*^, i.e. the slope of release, is established during model fitting during the first phase and is critical to establish a rise in cases. We utilized a best-fit model that optimized a loss function. After lockdown removal in June, a release of population that adopts the behaviour of the baseline is estimated. termed the second release. Using the baseline scenario we quantified the second release with respect to the release of population (given by the rate *κ*^*t*^) into the susceptible population during the first phase. We list the key features and parameters of our methodology below:

- Baseline rate of population increase (rate *κ*_*t*_) was established during February to March, 2021. An optimization method was used to achieve a best-fit model for the rise in confirmed cases and deaths.
- Average Vaccination rates: 3.0M/day or approximately 0.225%*/*day till August 16th.
- Subsequent rates averaged to 5M vaccinations/day. Optimistic projected vaccination during releases from lockdown is averaged to approximately 8M vaccinations/day for first dose.
- Additional lockdown removal date is estimated in late November due to local cultural events and its impact estimated to be starting on December 16th, 2021. Lockdown release rate are 100% of baseline over 30 days (high release) and 60 days (moderate release) given the lack of social distancing during the months of November and December.
- We modeled the loss of efficacy of vaccines as reduced linearly over 4 months by 50%. This loss of efficiency results in breakthrough infections.
- The efficacy of vaccines against Omicron was incorporated, considering two case with 50% and 75% of the original efficacy.
- We assume that there is a seeding of 50 cases per state on December 15th, 2021.

We provide two scenarios, one where there will be (i) no subsequent lockdowns and (ii) there will be mild lockdowns, impacting the social distancing and the susceptible population by a factor of 15, 20 and 25 %.

## Discussion

The modeling shows projections (Figure 3) that were concerning, especially due to reduction in social distancing countered by reported milder symptoms of the new variant. We modeled this release in two scenarios where the susceptible population is increasing at a substantial rate, the rate being represented by a release of the estimated base population in 30 and 45 days. The first corresponds to a fast release and the second to a moderate release. The estimates illustrate mixed impact of the two variants, with the Omicron variant rapidly dominating the currently existing Delta variant.

**Figure 3:**
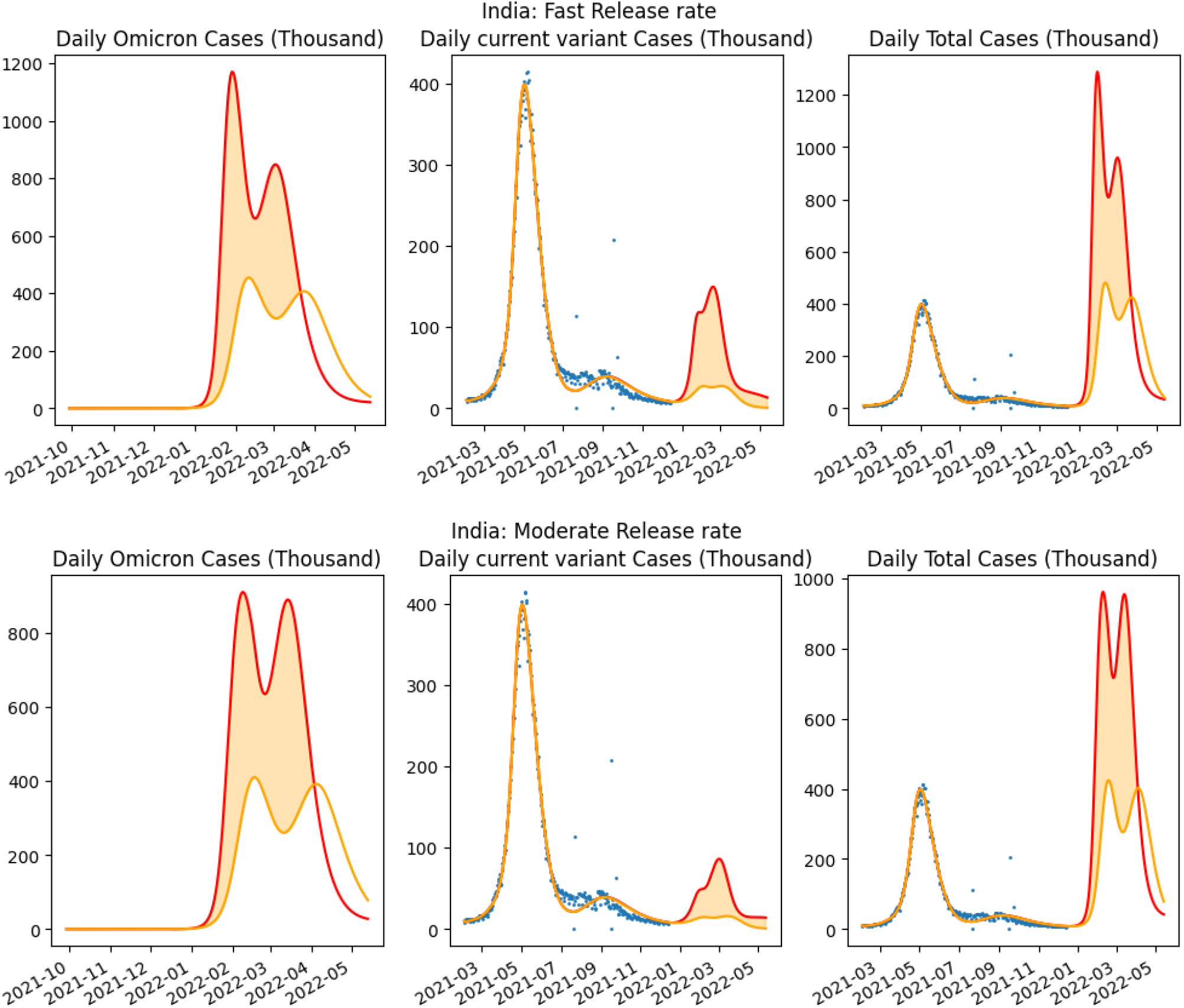
50% Vaccine efficacy against Omicron, No lockdown: Comparison of two scenarios: releasing population having occurred from lockdown illustrating two release rates, high (30 days) and moderate (60 days) Red indicates the worst case behavior (Scenario 1) and Orange indicate the optimistic Scenario 2. The leftmost graph in each row is for the projected cases of the omicron variant whereas the third graph is for the projected confirmed cases of the two variants together.

We determined estimates from our parameterized model when there was no lockdown as compared to the situation when there was partial lockdown initiated.

### No Lockdown assumption

The two scenarios considered are provided below and evaluated in December, 2021. The indicated peaks for daily infections are substantial.

#### Scenario 1 (pessimistic scenario–red curve)

The pessimistic scenario is when the numer of future release into susceptible pool is adopted to be equivalent to the release during the established baseline estimate. The population that has already become part of the susceptible pool after the relaxation from lockdown is ignored.

This results indicated a substantial rise in infections when rules regarding lockdown are relaxed. Our results projected that there would be increasing high daily case loads peaking at the end of January, 2022 and the beginning of February 2022, extending into March and April months. This indicated that utmost care be taken to ensure social distancing and mask usage, reduced capacity of interior locations and marketplaces, especially in restaurants and bars [25]. Our model indicated infection cases peaking at around 1.3 Million cases per day for 50% efficacy loss and 1.0 Million cases for 25% efficacy loss.

Further peaks were estimated to be progressively delayed and limited in size if social distancing measure are strictly followed. Note that this does not account for any variants that can breakthrough the vaccine defense. The second wave of infections in March to June 2021 illustrated the impact of lack of continued emphasis on these measures.

#### Scenario 2 (optimistic scenario-orange curve)

This scenario assumes that the susceptible that are added after relaxation of lockdown, but before 15th December, 2021, as estimated from the release function *κ*^*t*^ = *h*^*R*^ are discounted from future release of population into the susceptible pool (supplementary table S7). This is an optimistic scenario which models reduced susceptible size due to spreading infection. Even then this indicated a substantial rise in daily case numbers peaking at above 400 thousand cases.

These two scenarios are illustrated in Figure 3 and Figure 4.

**Figure 4:**
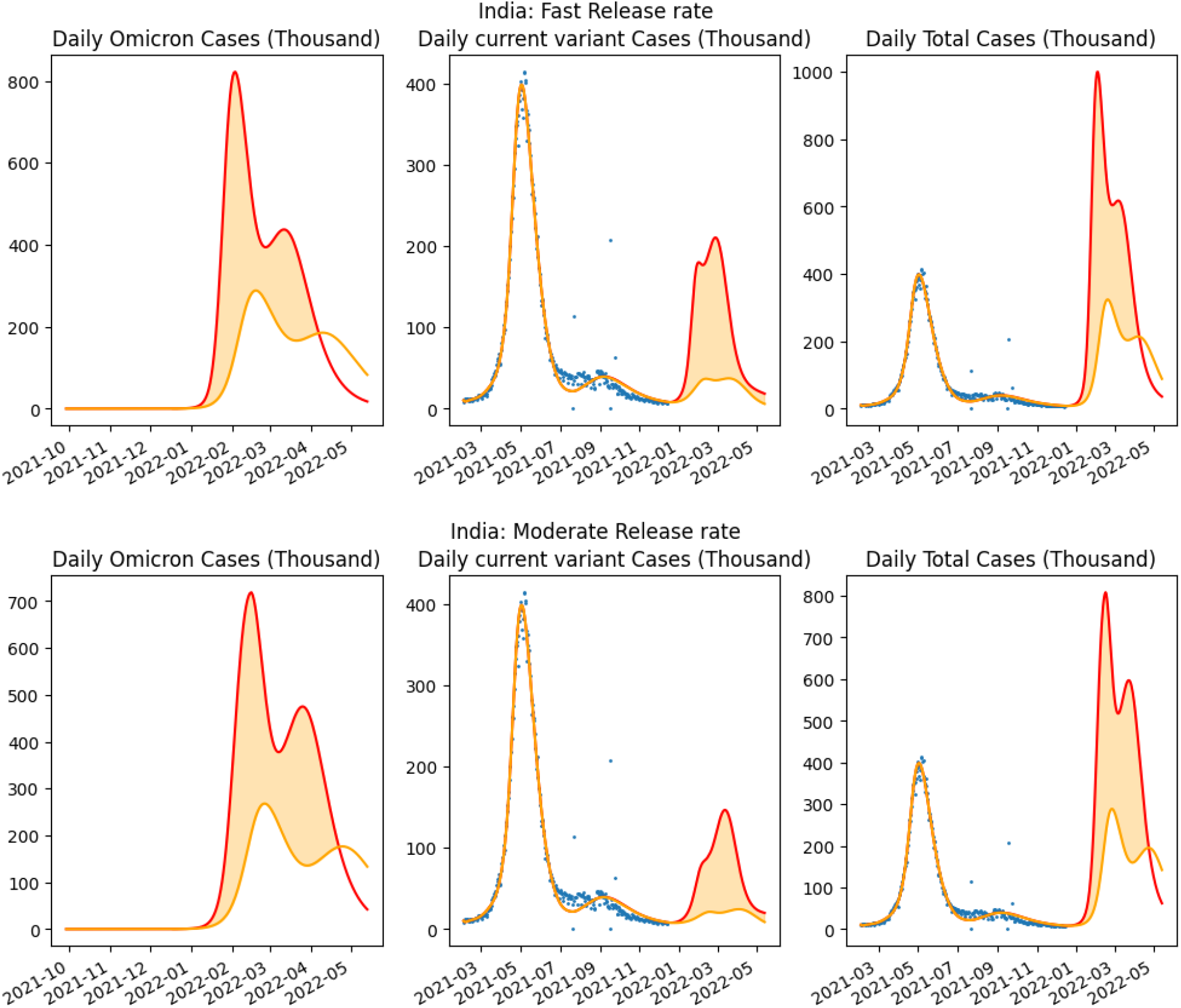
75% Vaccine efficacy against Omicron, No lockdown: Comparison of two scenarios: releasing population having occurred from lockdown with high release (30 days) and low release (60 days). Red indicates the worst case behavior (Scenario 1) and Orange indicate the optimistic Scenario 2. The leftmost graph in each row is for the projected cases of the omicron variant whereas the third graph is for the projected confirmed cases of the two variants together.

### Impact of Lockdown

We also consider the situation when a mild lockdown is implemented after the first week of January, when substantial cases will be visible. January 8th was chosen as the lockdown date in our simulation. This lockdown is assumed to reduce the susceptible population by 15, 20 and 25% for the high release rate scenario (30 day release) prior to the date of lockdown. It is assumed that the release of population stops, and is actually reversed, as the lockdown is implemented.

We observe substantial benefits due to partial lockdown, being limited to less than half of the cases at peak when no lockdown is enforced (Figures 5 and 6), which would have resulted in substantial cases and the resulting hospitalization could have impacted the health system.

**Figure 5:**
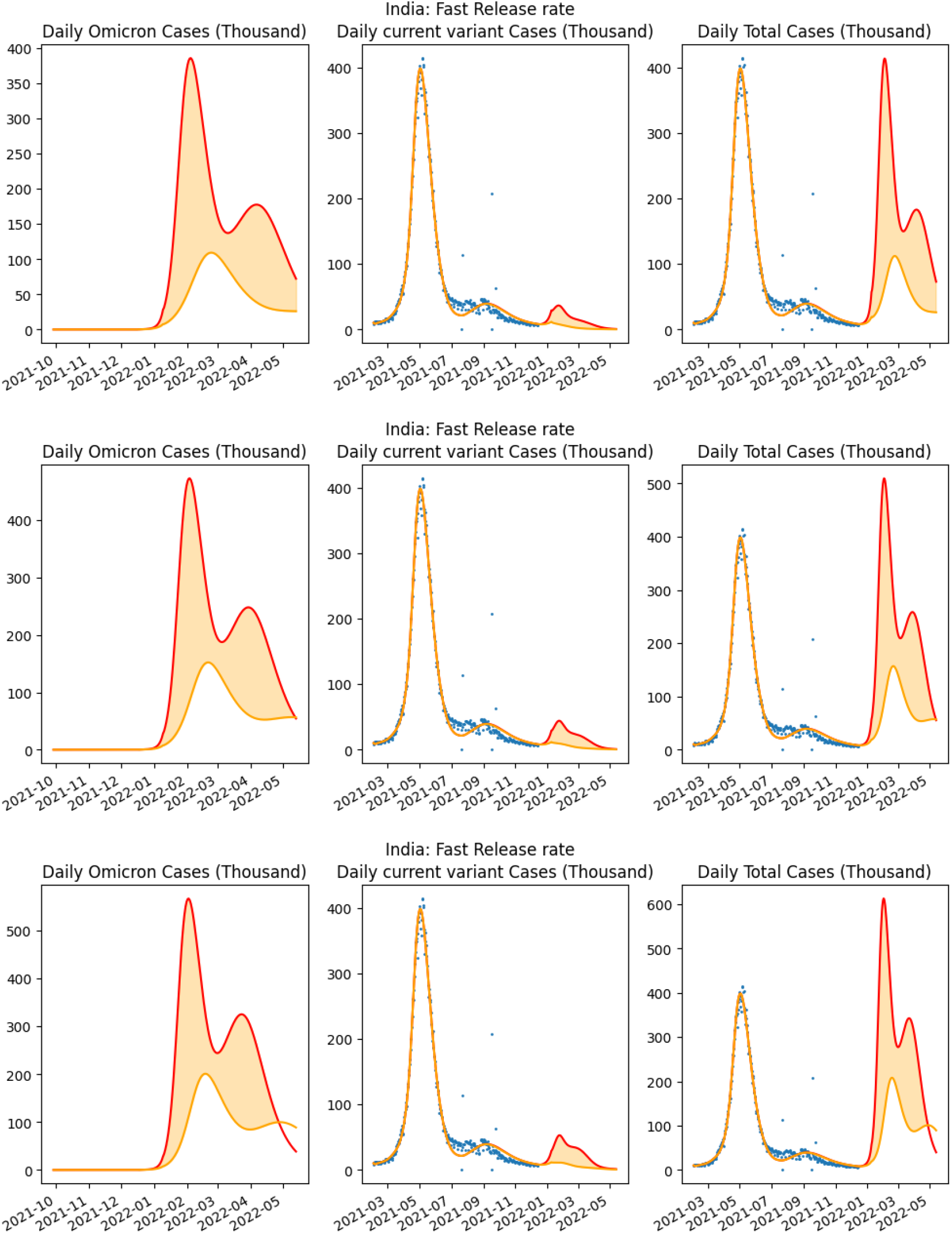
50% Vaccine efficacy against Omicron with Lockdown proportion 25,20 and 15 % startingJanuary 8th: Comparison of three scenarios based on the proportion of population under lockdown. The leftmost graph in each row is for the projected cases of the omicron variant whereas the third graph is for the projected confirmed cases of the two variants together.

**Figure 6:**
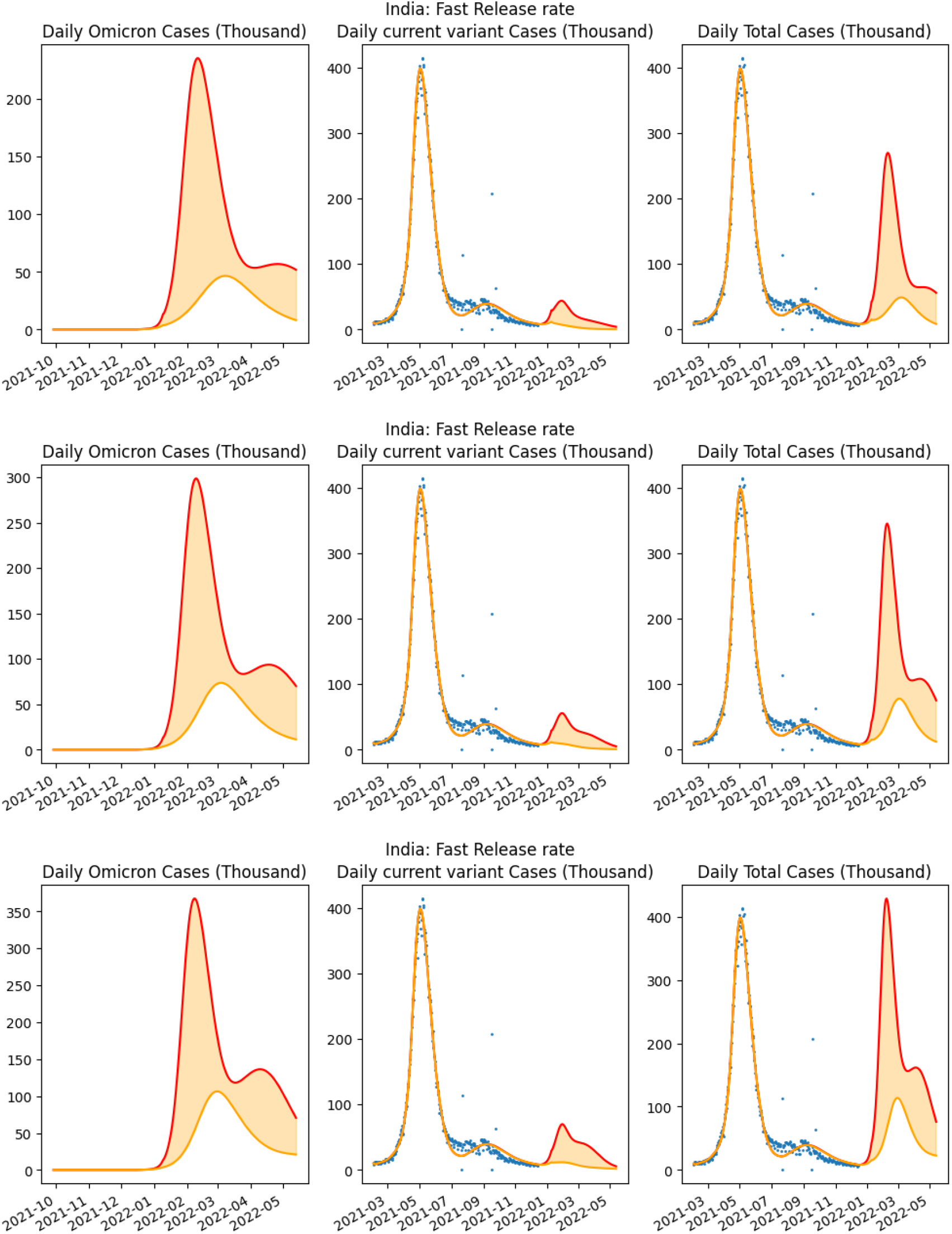
75% Vaccine efficacy against Omicron with Lockdown proportion 25,20 and 15 % startingJanuary 8th: Comparison of three scenarios based on the proportion of population under lockdown. The leftmost graph in each row is for the projected cases of the omicron variant whereas the third graph is for the projected confirmed cases of the two variants together.

The benefits of partial lockdown are further illustrated in figure 7 where the peak number of confirmed cases shows an almost linear decrease with respect to the increase in the percentage of people in partial lockdown. The results indicate that partial lockdown of 25% is reasonably effective in bringing the case incidence down by about 50% even with low vaccine efficacy (Efficacy=0.2). Furthermore, figure 8 shows that delaying partial lockdown increases the possible peak daily infections. It was indicated to be critical that partial lockdown be implemented as soon as feasible.

**Figure 7:**
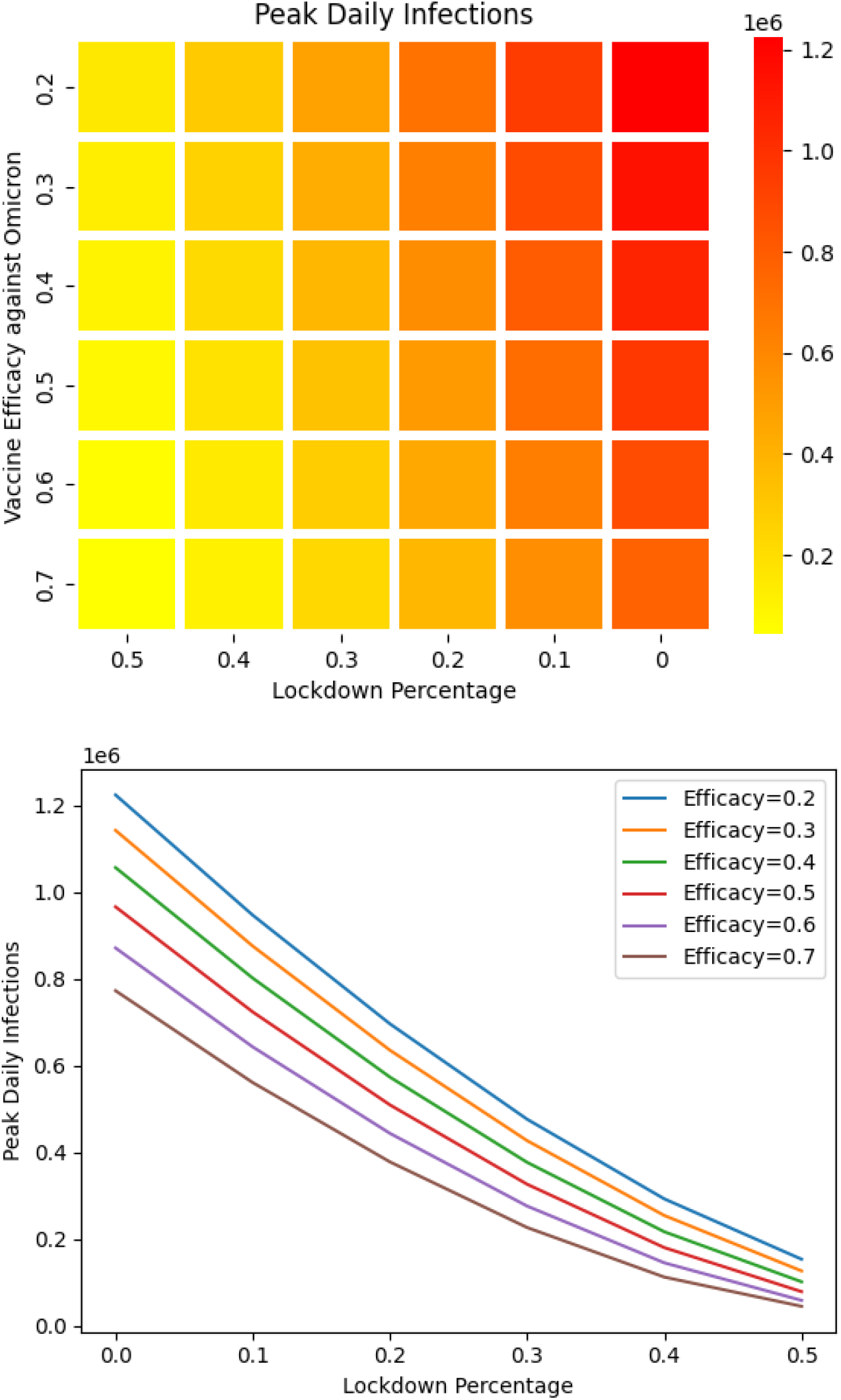
Predicted Peak of confirmed cases as dependent on efficacy of vaccine against the Omicron variant and percentage of population under lockdown. These illustrations are for the high release rate scenario. The release stops at the time of lockdown.

**Figure 8:**
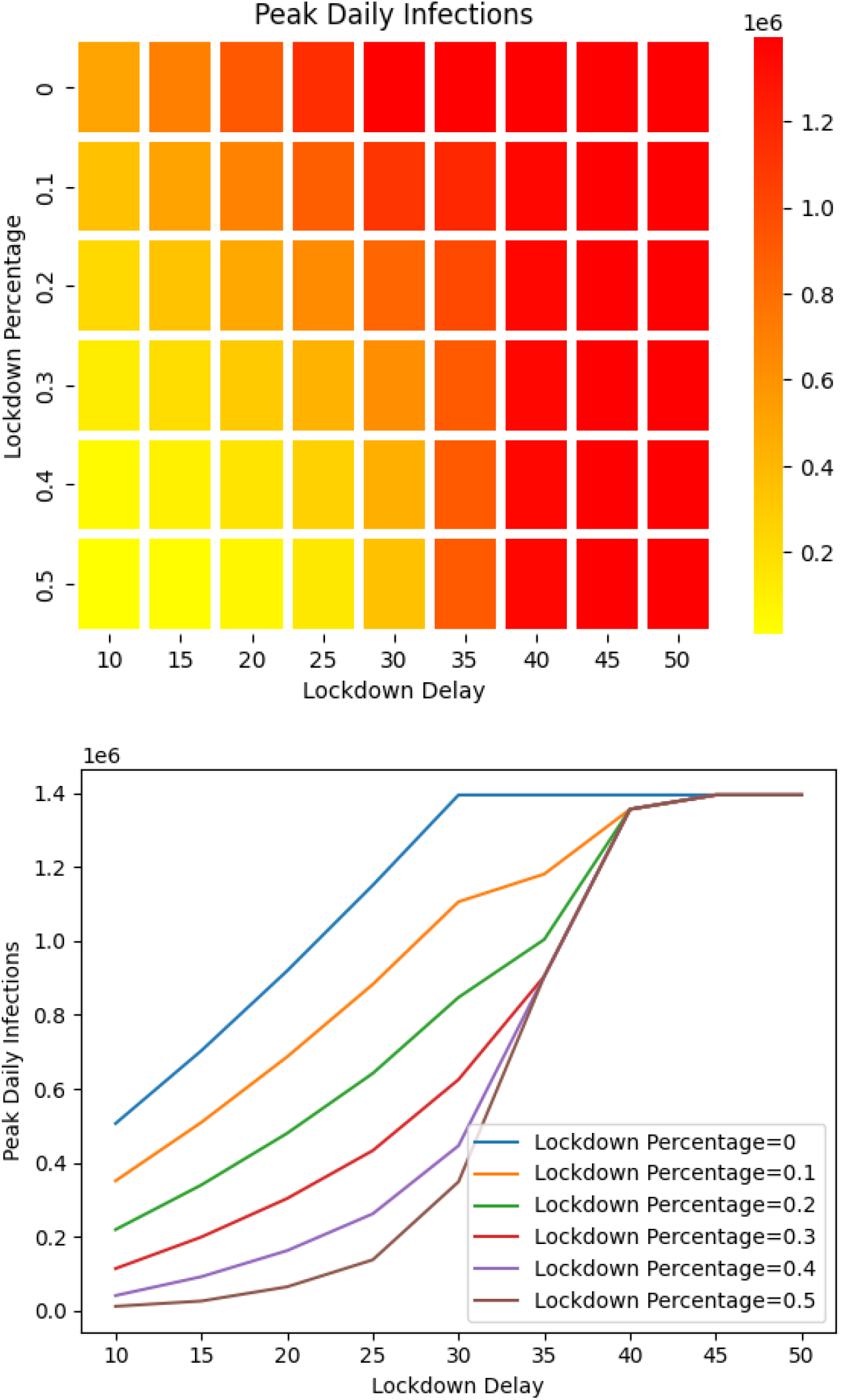
Predicted Peak of confirmed cases as dependent on percentage of population under lockdown and delay of lockdown. These illustrations are for the high release rate scenario. The release stops at the time of lockdown.

## 4 Methodology

Model parameters were determined by an optimization method to minimize a loss function that is dependent on cumulative cases and deaths.

We let *G, D* denote the simulated cumulative cases and deaths, 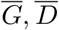 are the corresponding reported data. Our optimization methodology utilizes a random initial choices of the parameters over defined ranges as well as a search over the space of release dates to achieve the best fit. The ranges were chosen to be wide enough so that the optimum point was not constrained. The optimization method used is L-BGFS that converges to a best-fit estimate of the model parameters, which due to the non-convex nature of the problem could possibly be a local optima. Sampling multiple starting points to find the best fit leads to reasonably accurate fitting as measured by *R*^2^ values.

### 4.1 First-Phase fitting

In the first phase we determined the parameters related to the first variant of the virus. We consider the period from February up till June 10th, 2021 when the removal of lockdown began, as an effective release phase where the population behavior trended towards low mask usage and high mobility. While around mid April, 2021 there was a lockdown, we consider the entire period as an effective release period. The release of susceptible population was assumed to be a linear function with a starting date in a range of 1 week beginning on 15th March. These dates were sampled to compute the release function that resulted in the best fit.

The parameters we considered for fitting are *β, γ*_*E*_, *α, γ, γ*_2_, *γ*_3_, *a*_1_, *a*_2_, *a*_3_, *h*_*R*_, *t*_1_, *t*_2_ that are used to generate the series *G* and *D* according to the differential equations governing the model.

We use the following loss function.

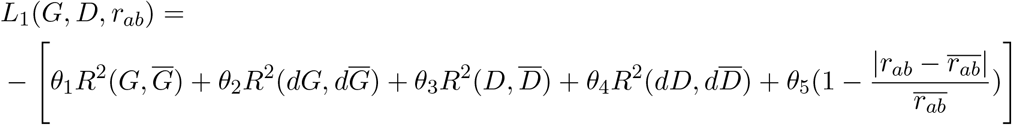

where the series *G, D* and 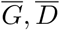 are weighted geometrically emphasizing the latest data points in each phase, *R*^2^ is the coefficient of determination, 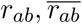 are the antibody ratio at the end of the first phase of the simulation and reported, respectively. We obtained 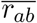 from sero-surveys carried out by ICMR [17, 20] Our simulation is on a daily basis with 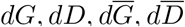 being calculated daily. Relative weighting of the parameters *θ*_*i*_, *i* = 1, 2, 3, 4, 5 was used to fit the multi-objective function, the choices being provided in the supplementary material.

### 4.2 Second-Phase Fitting for reopen

In the second phase when removal of lockdown started we considered release of people from lockdown via a linear function with a starting date in a range after 1 week of release of lockdown, i.e. starting on the 17th of June. The release of population into the susceptible set is assumed to be a linear function again and the rate of release dictates the speed at which the policy restrictions are being relaxed and the behavior of the population.

The growth of the infection curve is strongly dependent on this rate, as we will illustrate in our projections. During this phase we estimate the rate of release, the start of release and the size of the release.

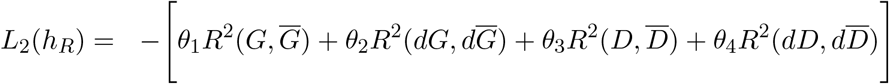

### 4.3 Fitting parameters and Projection accuracy

We illustrate our fitting for a combined first and second phase in Figure 9. The goodness of fit for all Indian states is illustrated in the supplementary material. The *R*^2^ values illustrating the goodness of fit was determined and listed in Table S4 in the supplementary material. The box plot in Figure 9 illustrates that the per-capita RMSE is bounded to be less than a small fraction with maximum at less than 0.1% indicating a high degree of accuracy, with a corresponding range of *R*^2^ scores close to 1.

**Figure 9:**
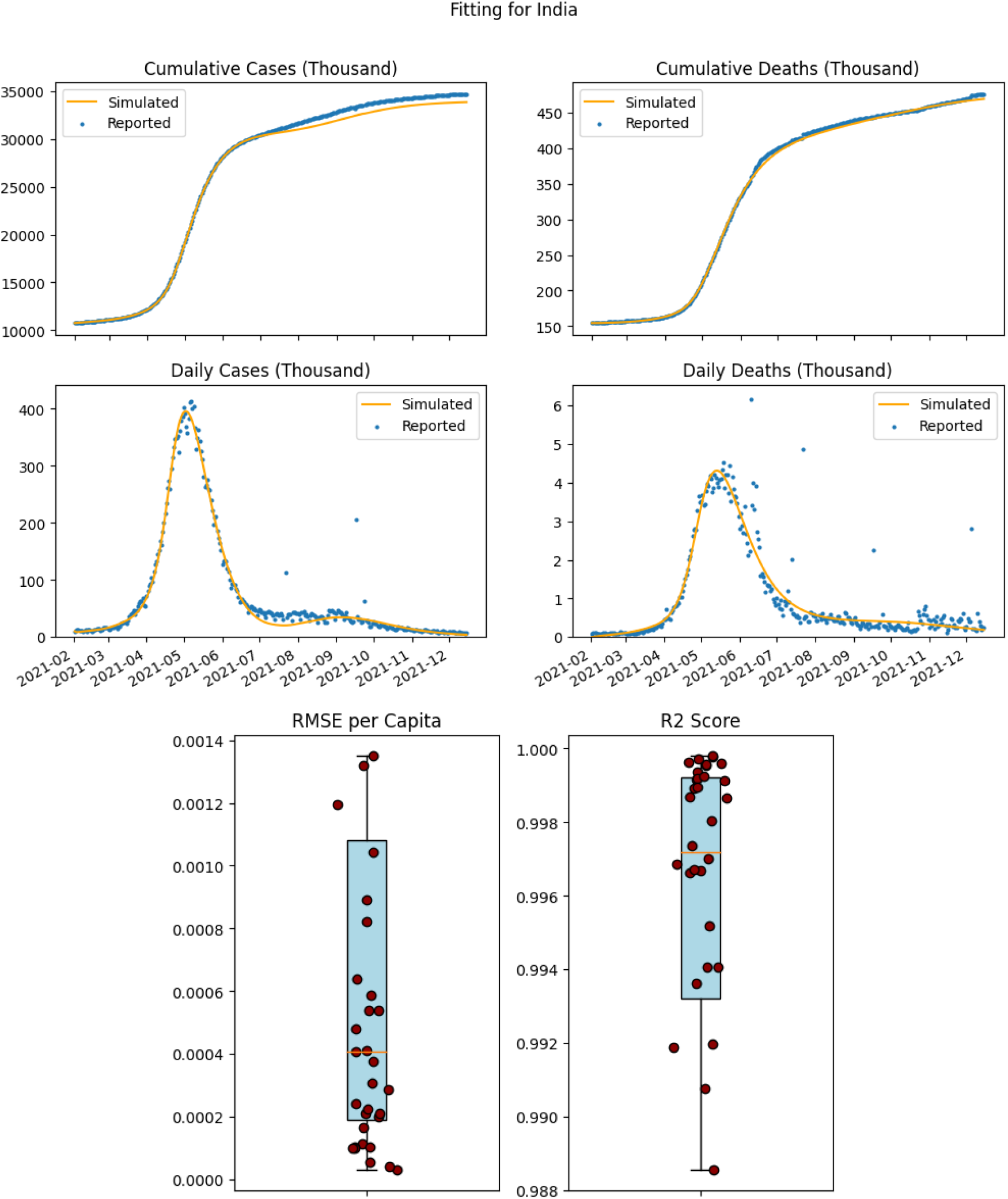
Fitting results for India, obtained by accumulating fitting for each state within India, illustrates model performance for both confirmed cases and deaths during the period from February 1st to 15th December, 2021. Data indicating cumulative numbers for each state was used and the model’s performance in terms of the per capita RMSE and *R*^2^ scores for each of the states shows the accuracy of the model. Outliers were removed from the Box-plots. Oulier data is in the supplementary material **??**

To determine the accuracy of our projections, before the start of the projections, we estimated parameters for our model using data until May 11th, 2021. We fit parameters for the first phase of our model using 4 sets of experiments where we used training data until 4/11/2021, 4/18/2021,4/24/2021, 4/30/2021 and estimated the error in projections of daily cases of infections and deaths beyond these dates for 1-4 weeks. The error in projections of daily cases beyond the 4 chosen training dates was below 5% as detailed in the supplementary material S2. Since the vaccination rates were still low, the projections used for testing the accuracy of the model did not account for vaccinations.

### Projection accuracy for the Omicron Wave

The projection accuracy was measured by evaluating RMSE, MAPE and *R*^2^ values comparing the projections and the actual confirmed cases. We compared the error in our pessimistic estimates versus the actual data for cumulative and daily confirmed cases and deaths at three time intervals, corresponding to 15, 30 and 45 days starting from 15th December, 2021. Cumulative cases were counted from December 15th, 2021. For cumulative cases, the RMSE error per capita was extremely small for projections 15-45 days forward with the maximum error being 0.0013 per capita. The *R*^2^ values were 0.744,0.36,0.203 for 15, 30 and 45 day projections, respectively. And MAPE measure was 17.8%, 27.2% and 41.6% for 15,30 and 45 day intervals respectively. The corresponding numbers for cumulative deaths were: *R*^2^ values of 0.74,0.803 and 0.936 for the 15, 30 and 45 day interval projections. The MAPE values were 20.9%, 21% and 16.9%, while RMSE error/per capita was negligible. For daily cases, there was more variability and the details of the errors may be found in Table S3.

Our projections were based on initial estimate of 50 Omicron cases on December 15th, 2021, in each state, chosen due to the absence of confirmed numbers as accurate Omicron variant case numbers were unknown at that stage.

## 5 Conclusions and Limitations

Early warning systems which project long range behavior more than a month in advance, while not able to pinpoint exact peaks and accurate case projections, are very useful for policy decisions. Our results were made available to a nodal body (at the central government) for policy decisions. Partial lockdowns, based on inputs from multiple sources, were implemented by various state governments in India.

As indicated from the results, which re-evaluated projections monthly, long term projection of the disease is impacted by governmental policy decisions and adherence to social distancing policies by the population. While it appears that policy decisions, including vaccinations, induced monthly shifts delaying the third wave until December 2021, further investigation of the impact of the policies is required. While we have accounted for a measured reduction of lockdown as well as investigated implementation of levels of lockdown strategies, the impact of a variety of factors including adherence to policy declarations remains an important aspect of further study.

We would like to thank Dr. V.K. Paul (Niti Aayog, Government of India) and Professor S.N. Maheshwari (Honorary Professor, IIT Delhi) for their suggestions and help during the development of these projections.

## Data Availability

Data is available on repositories

## Supplementary Materials

### S1 *R*_0_ with multiple variants

We determine the value of *R*_0_ for a SEIR model with multiple virus variants comprising the set *C*. The equations governing the behavior of this system are as follows:

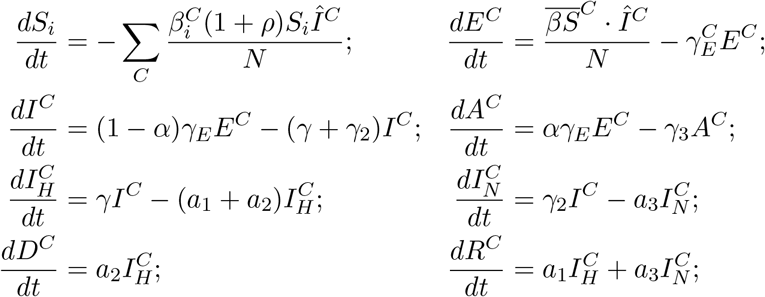

where *ρ* is the rate at which the susceptible set changes due to the population during relaxation of lock-down or during lockdown 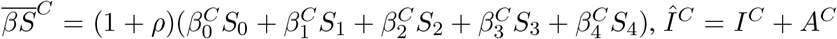. The parameter *ρ* is negative in a lockdown state and positive when lockdown is released. The growth *R*_0_ can be obtained from the simplified linear subsystem, where *S* = *S*_0_ + *S*_1_ + *S*_2_ + *S*_3_ + *S*_4_ is assumed to be *N* :

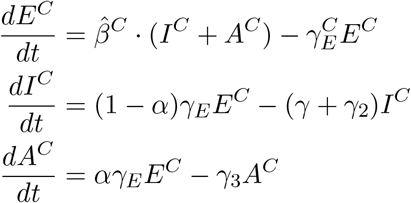

where 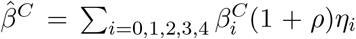 for the types of the variant. The fractions *η*_*i*_ represent the distribution of susceptibles into the various groups *S*_0_, *S*_1_, *S*_2_, *S*_3_ and *S*_4_. Using the method of [8] we can estimate the value of *R*_0_ from the eigenvalues of *T* Σ^−1^ where *T* and Σ are block diagonal matrices: 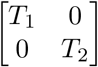 and 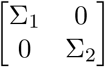 with

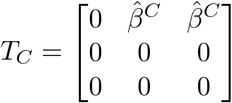

and

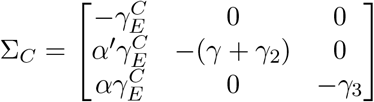

where *α*^*′*^ = 1 − *α*. Thus

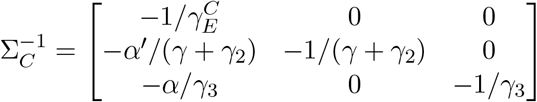

The dominant eigenvalue of *T* Σ^−1^ is 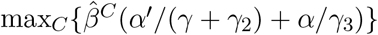

### S2 Model and Projection Accuracy

To establish the accuracy of our model we estimated parameters for our model using training data and projected until May 11th, 2021. We trained our our model using data until 4/11/2021, 4/18/2021,4/24/2021, 4/30/2021 and estimated the error in projections of daily cases of infections and deaths. The error for projections beyond the 4 chosen dates is illustrated in Table S1 and Table S2. Since death projections had a time lag, projections were limited as the training data dates progressed.

**Supplementary Table S1:** Error in daily new case projections (all India)

| Fitting Date | 1 week | 2 weeks | 3 weeks | 4 weeks |
| --- | --- | --- | --- | --- |
| 4/11/21 | 4.54% | 4.11% | 3.85% | 4.13% |
| 4/18/21 | 3.67% | 3.51% | 3.99% |  |
| 4/24/21 | 3.41% | 4.21% | 4.4% |  |
| 4/30/21 | 4.37% | 4.7% | 5.0% |  |

**Supplementary Table S2.** Error in daily new death projections (all India)

| Fitting Date | 1 week | 2 weeks | 3 weeks | 4 weeks |
| --- | --- | --- | --- | --- |
| 4/11/21 | 11.15% | 7.48% | 7.73% | 6.83% |
| 4/18/21 | 3.80% | 6.02% | 5.39% |  |
| 4/24/21 | 8.64% | 6.42% |  |  |
| 4/30/21 | 5.33% |  |  |  |

#### Omicron Projection Accuracy

We measured the accuracy of our long range projections during the Omicron phase. We measured the cumulative and daily case count errors at three intervals, 15, 30 and 45 days after December 15th, 2021. The results are shown in the Table S3.

**Supplementary Table S3.** Error in Case Projection of Omicron wave starting from 12/15/2021

| metric | 15 days | 30 days | 45 days |
| --- | --- | --- | --- |
| RMSE per capita(daily cases) | 0.00000250 | 0.00003963 | 0.00027739 |
| R2(daily cases) | -12.0704 | 0.5361 | -6.1591 |
| MAPE(daily cases) | 38.115% | 40.541% | 80.549% |
| RMSE per capita(daily death) | 0.00000010 | 0.00000011 | 0.00000015 |
| R2(daily death) | -0.8340 | -0.1965 | -0.5837 |
| MAPE(daily death) | 32.621% | 37.510% | 39.884% |
| RMSE per capita(cumulative cases) | 0.00001232 | 0.00028131 | 0.00137607 |
| R2(cumulative cases) | 0.7440 | 0.3763 | 0.2032 |
| MAPE(cumulative cases) | 17.857% | 27.268% | 41.611% |
| RMSE per capita(cumulative deaths) | 0.00000059 | 0.00000095 | 0.00000093 |
| R2(cumulative deaths) | 0.7405 | 0.8036 | 0.9360 |
| MAPE(cumulative deaths) | 20.989% | 21.112% | 16.869% |

### S3 Results of model fitting for States in India

The results for the fitting model fitting are illustrated in Figure S2. The fitting was over two phase, the first phase ending on June 10th, the lockdown phase and the second phase was the release phase from June 10th on-wards till December 15th.

The *R*^2^ values for the combined fitting, i.e. first and second phase, of the confirmed infection case numbers is contained in Table S4 in the first column while the *R*^2^ values for the combined fitting of the number of deaths is in the 4th column. The *R*^2^ values were consistently high over all the states. *R*^2^ values for individual states are inconsistent in the second phase for both the infected cases and the deaths since each individual state had applied varying policies, some including multiple attempts at restrictive policies including lockdown during this phase. The parameters obtained from the fitting are detailed in Table S6.

**Supplementary Table S4.** R^2^ score for India and all states resulting from fitting the model

| State | $R^2$(confirmed) | $R^2$(confirmed phase1) | $R^2$(confirmed phase2) | $R^2$(death) | $R^2$(death phase1) | $R^2$(death phase2) |
| --- | --- | --- | --- | --- | --- | --- |
| India | 0.9961 | 0.9997 | 0.7951 | 0.9993 | 0.9998 | 0.9750 |
| Andaman and Nicobar | 0.9967 | 0.9962 | 0.5327 | 0.1906 | 0.9662 | -1831.9827 |
| Andhra Pradesh | 0.9970 | 0.9999 | 0.7953 | 0.9504 | 0.9899 | -0.3936 |
| Arunachal Pradesh | 0.9952 | 0.9980 | 0.9640 | 0.9925 | 0.9725 | 0.9446 |
| Assam | 0.9379 | 0.9998 | -0.4587 | 0.9603 | 0.9977 | 0.2740 |
| Bihar | 0.9997 | 0.9996 | -0.6258 | 0.0798 | 0.9389 | -63.8096 |
| Chandigarh | 0.9987 | 0.9982 | 0.8754 | 0.9540 | 0.9930 | -0.4292 |
| Chhattisgarh | 0.9996 | 0.9997 | -0.3986 | 0.9994 | 0.9995 | -1.3777 |
| Daman and Diu | 0.9996 | 0.9996 | -0.0732 | 0.6843 | 0.6575 | 0.0000 |
| Delhi | 0.9996 | 0.9996 | -4.5287 | 0.9996 | 0.9991 | -0.4389 |
| Goa | 0.9908 | 0.9988 | -0.6514 | 0.9859 | 0.9991 | -0.3780 |
| Gujarat | 0.9998 | 0.9997 | -1.8440 | 0.9979 | 0.9934 | 0.6432 |
| Haryana | 0.9994 | 0.9981 | -0.0964 | 0.9795 | 0.9993 | -2.6716 |
| Himachal Pradesh | 0.9886 | 0.9931 | 0.1555 | 0.9966 | 0.9910 | 0.7871 |
| Jammu and Kashmir | 0.9941 | 0.9991 | -0.1017 | 0.9954 | 0.9943 | -0.4672 |
| Jharkhand | 0.9998 | 0.9995 | 0.7872 | 0.9927 | 0.9984 | -187.2522 |
| Karnataka | 0.9991 | 0.9999 | 0.7588 | 0.9989 | 0.9995 | 0.8907 |
| Kerala | 0.9633 | 0.9997 | 0.8058 | 0.9825 | 0.9821 | 0.9538 |
| Ladakh | 0.9941 | 0.9955 | -0.6089 | 0.7665 | 0.9558 | -55.2289 |
| Lakshdweep | 0.9223 | 0.9985 | -8.3217 | 0.7206 | 0.9570 | -35.6842 |
| Madhya Pradesh | 0.9989 | 0.9997 | -40.8878 | 0.7687 | 0.9852 | -6.0478 |
| Maharastra | 0.9919 | 0.9991 | 0.2586 | 0.9848 | 0.9975 | 0.5334 |
| Manipur | 0.9967 | 0.9972 | 0.9811 | 0.9472 | 0.9714 | 0.6193 |
| Meghalaya | 0.8741 | 0.9937 | 0.0778 | 0.9854 | 0.9900 | 0.8856 |
| Mizoram | 0.5215 | 0.9979 | 0.0192 | 0.9473 | 0.9899 | 0.8839 |
| Nagaland | 0.9974 | 0.9988 | 0.9628 | 0.9327 | 0.9913 | -0.0927 |
| Odisha | 0.9966 | 0.9998 | 0.8474 | 0.5614 | 0.9976 | -0.8083 |
| Puducherry | 0.9920 | 0.9977 | 0.2387 | 0.9932 | 0.9957 | -0.0703 |
| Punjab | 0.9989 | 0.9981 | -0.7284 | 0.9992 | 0.9989 | 0.7297 |
| Rajasthan | 0.9992 | 0.9988 | -33.4340 | 0.9990 | 0.9996 | -9.8509 |
| Sikkim | 0.9987 | 0.9989 | 0.9882 | 0.8127 | 0.9726 | -1.4990 |
| Tamil Nadu | 0.9992 | 0.9976 | 0.9520 | 0.9431 | 0.9974 | -2.1433 |
| Telangana | 0.9969 | 0.9995 | 0.7377 | 0.9963 | 0.9982 | 0.7271 |
| Tripura | 0.9936 | 0.9985 | 0.9132 | -0.5002 | 0.9864 | -23.2177 |
| Uttar Pradesh | 0.9980 | 0.9998 | -158.6117 | 0.9873 | 0.9934 | -8.8216 |
| Uttarakhand | 0.9993 | 0.9984 | -0.4632 | 0.9972 | 0.9993 | -0.8500 |
| West Bengal | 0.9995 | 0.9996 | 0.9377 | 0.9911 | 0.9980 | 0.5844 |

**Supplementary Table S5.**
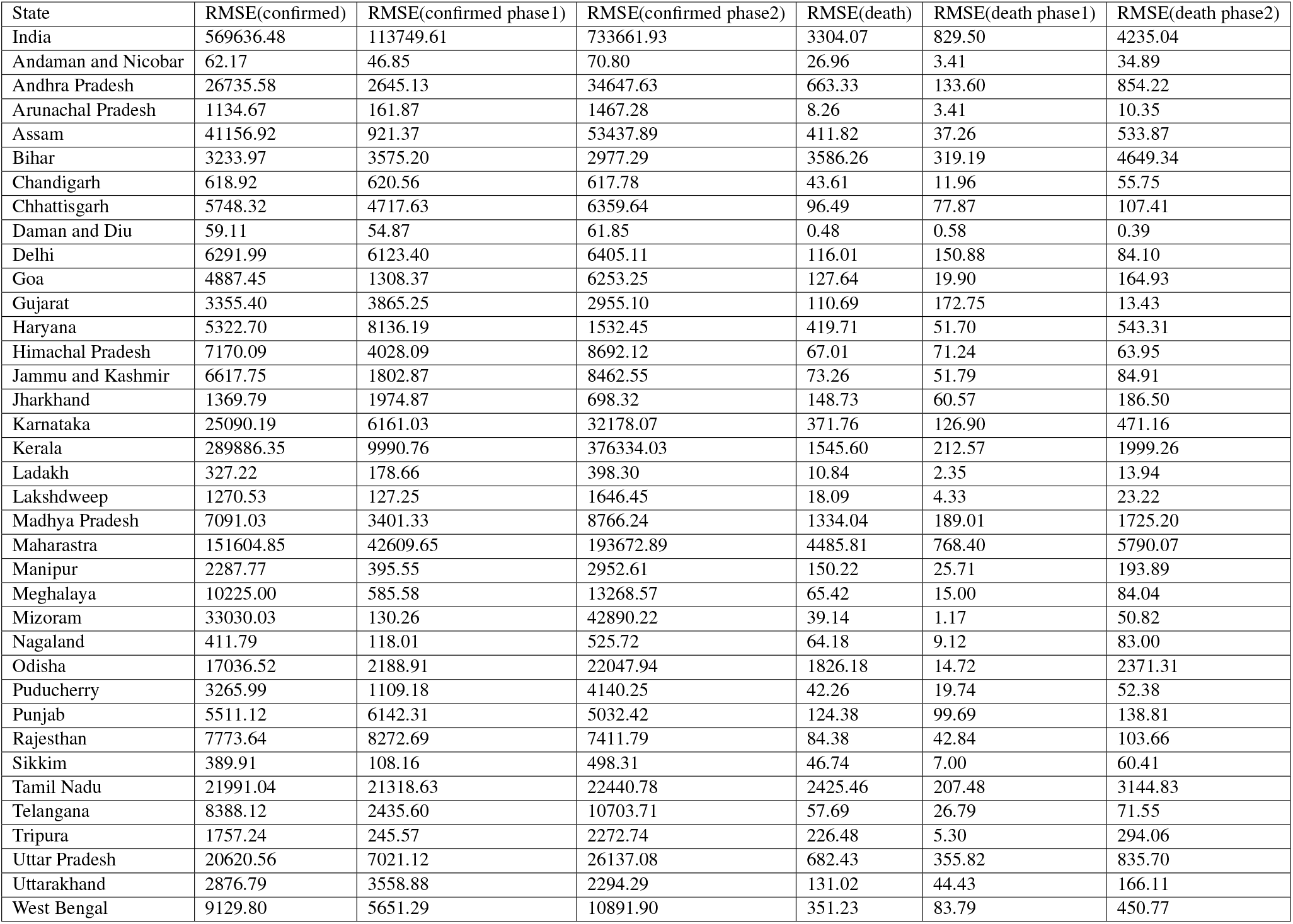
RMSE score for India and all states resulting from fitting the model

**Supplementary Figure S1.**
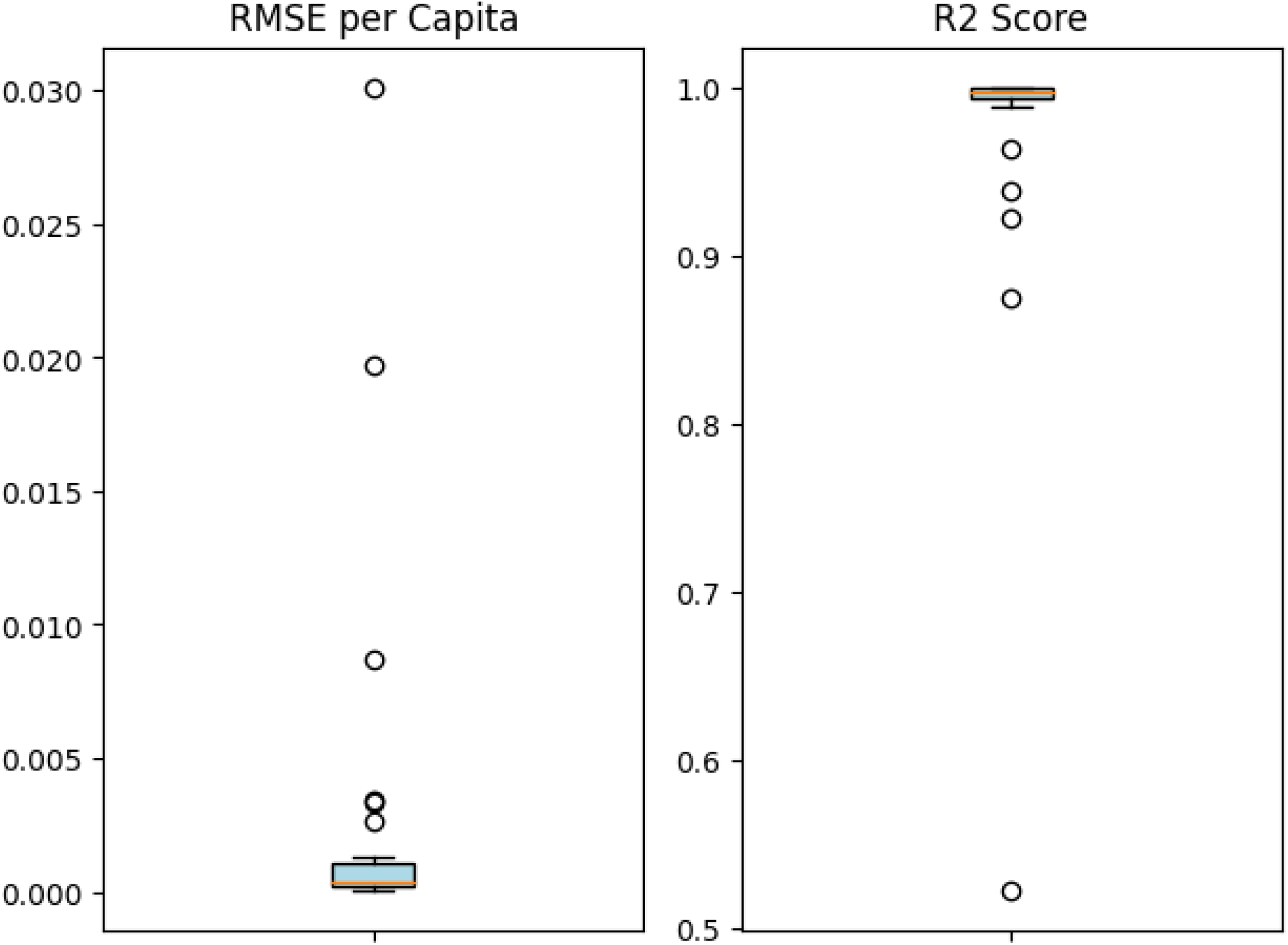
RMSE per Capita and R2 scores

**Supplementary Figure S2.**
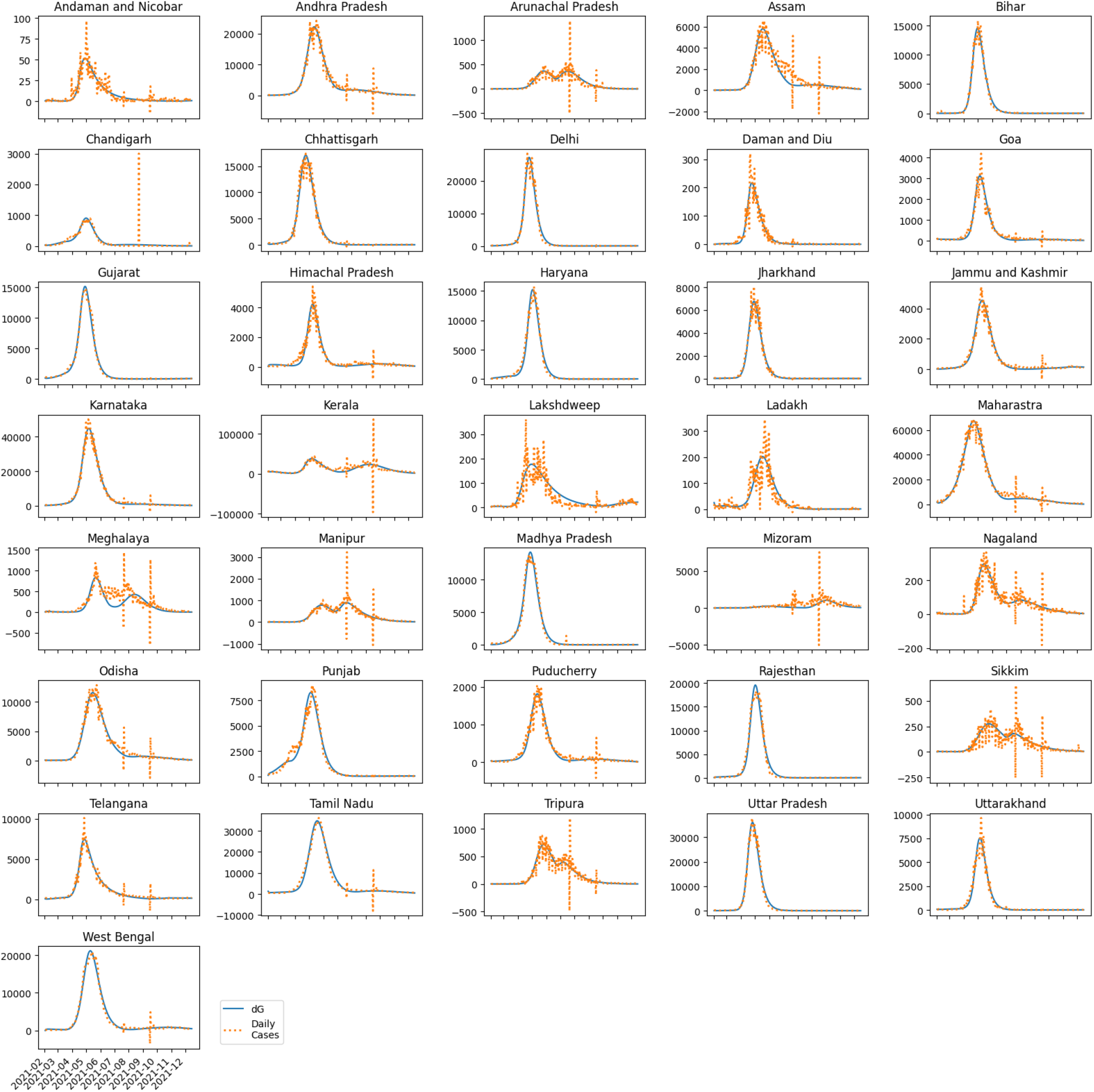
Fitting of Indian States

**Supplementary Figure S3.**
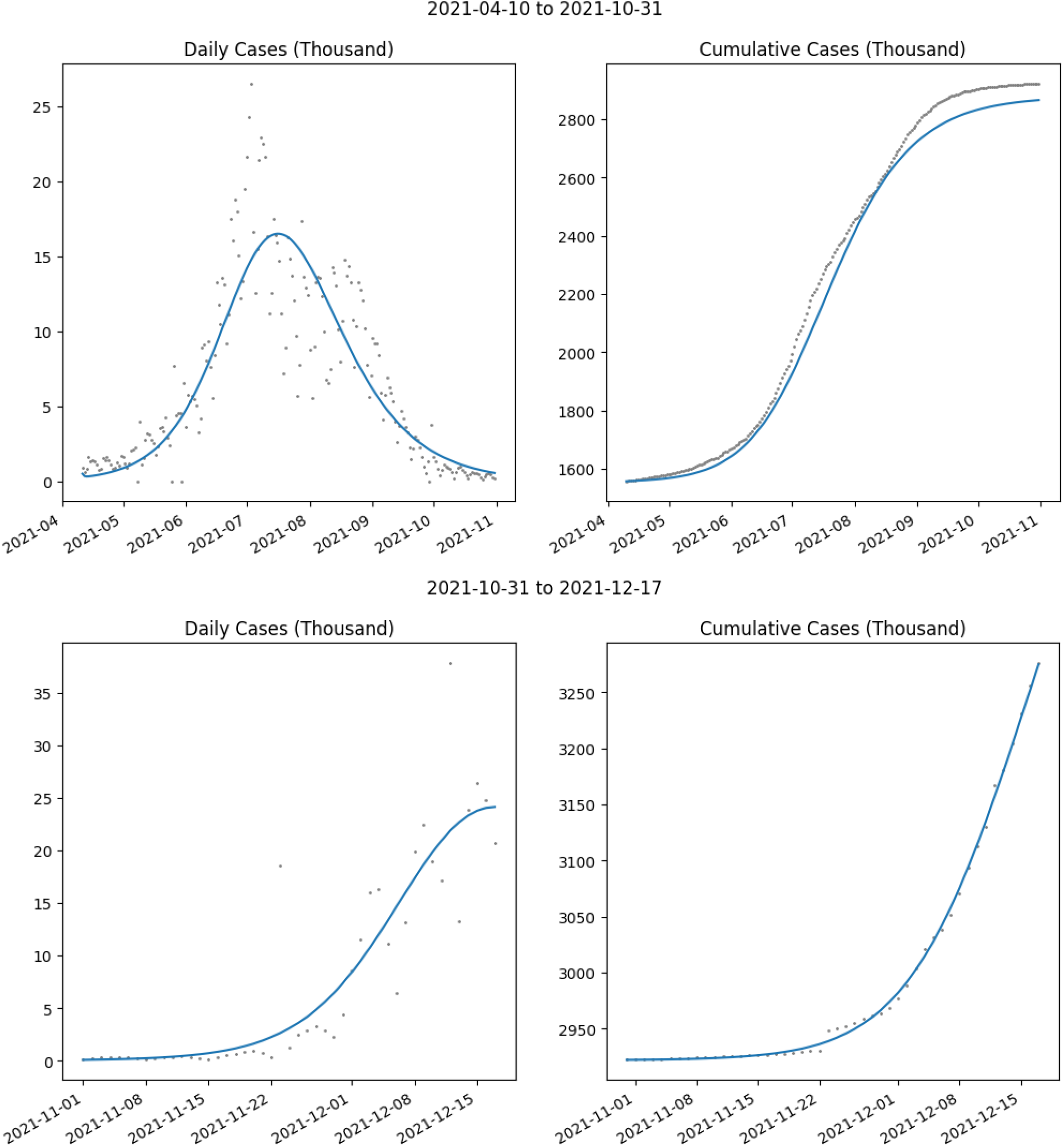
Fitting of South Africa. The ratio of *β* in the 2nd period versus the 1st is 1.8.

**Supplementary Table S6.** Parameters determined for all the states. The model was fit from February, 2021 to June 10, 2021.

| State | $\beta$ | $\gamma_E$ | $\alpha$ | $\gamma$ | $\gamma_2$ | $\gamma_3$ | $a_1$ | $a_2$ | $a_3$ |
| --- | --- | --- | --- | --- | --- | --- | --- | --- | --- |
| Andaman and Nicobar | 49.99956 | 0.3 | 0.275821 | 0.159024 | 0.060573 | 0.04 | 0.056469 | 0.003398 | 0.2 |
| Andhra Pradesh | 7.882238 | 0.295294 | 0.177333 | 0.040099 | 0.197636 | 0.199207 | 0.211944 | 0.007038 | 0.15097 |
| Arunachal Pradesh | 3.685567 | 0.2 | 0.779355 | 0.2 | 0.2 | 0.134107 | 0.5 | 0.005904 | 0.094896 |
| Assam | 4.621663 | 0.210916 | 0.815336 | 0.2 | 0.2 | 0.128884 | 0.5 | 0.013201 | 0.011474 |
| Bihar | 50 | 0.3 | 0.469605 | 0.063626 | 0.2 | 0.2 | 0.01 | 0.001736 | 0.01 |
| Chandigarh | 10.22508 | 0.3 | 0.1 | 0.2 | 0.2 | 0.2 | 0.114719 | 0.002874 | 0.013961 |
| Chhattisgarh | 7.971897 | 0.3 | 0.189233 | 0.154885 | 0.184541 | 0.2 | 0.33631 | 0.010948 | 0.021935 |
| Daman and Diu | 10.82086 | 0.3 | 0.870973 | 0.04 | 0.2 | 0.170437 | 0.5 | 0.001 | 0.165116 |
| Delhi | 10.7955 | 0.3 | 0.1 | 0.110269 | 0.2 | 0.2 | 0.137628 | 0.007119 | 0.01 |
| Goa | 5.666019 | 0.3 | 0.1 | 0.2 | 0.2 | 0.106537 | 0.222253 | 0.009409 | 0.015194 |
| Gujarat | 42.40642 | 0.3 | 0.1 | 0.2 | 0.2 | 0.2 | 0.5 | 0.010229 | 0.010791 |
| Haryana | 23.97823 | 0.3 | 0.1 | 0.2 | 0.2 | 0.2 | 0.118534 | 0.003017 | 0.011975 |
| Himachal Pradesh | 22.01681 | 0.3 | 0.1 | 0.2 | 0.2 | 0.2 | 0.235399 | 0.008519 | 0.013447 |
| Jammu and Kashmir | 14.32116 | 0.3 | 0.130402 | 0.2 | 0.199394 | 0.2 | 0.440694 | 0.011909 | 0.12049 |
| Jharkhand | 50 | 0.3 | 0.1 | 0.2 | 0.2 | 0.2 | 0.5 | 0.019294 | 0.015639 |
| Karnataka | 11.06724 | 0.210959 | 0.100545 | 0.045264 | 0.120618 | 0.199422 | 0.137694 | 0.006871 | 0.021345 |
| Kerala | 2.483146 | 0.21049 | 0.788614 | 0.04 | 0.2 | 0.112912 | 0.014261 | 0.001 | 0.099696 |
| Ladakh | 2.306169 | 0.295377 | 0.533661 | 0.06354 | 0.173535 | 0.199869 | 0.098133 | 0.002325 | 0.182724 |
| Lakshdweep | 1 | 0.2 | 0.369919 | 0.2 | 0.2 | 0.04 | 0.119597 | 0.001973 | 0.199669 |
| Madhya Pradesh | 49.99987 | 0.290687 | 0.107274 | 0.172474 | 0.2 | 0.2 | 0.086072 | 0.001673 | 0.013247 |
| Maharashtra | 4.339277 | 0.247925 | 0.156601 | 0.09567 | 0.142352 | 0.167112 | 0.027022 | 0.001429 | 0.2 |
| Manipur | 10.16117 | 0.2 | 0.370882 | 0.2 | 0.2 | 0.04 | 0.5 | 0.020669 | 0.2 |
| Meghalaya | 31.28439 | 0.296347 | 0.1 | 0.2 | 0.2 | 0.2 | 0.493665 | 0.023404 | 0.162305 |
| Mizoram | 1 | 0.2 | 0.898921 | 0.04 | 0.124766 | 0.064663 | 0.061328 | 0.002265 | 0.188491 |
| Nagaland | 28.61501 | 0.3 | 0.272491 | 0.056024 | 0.185681 | 0.061247 | 0.247969 | 0.046448 | 0.161904 |
| Odisha | 15.43327 | 0.206192 | 0.1 | 0.04 | 0.2 | 0.060363 | 0.03037 | 0.001 | 0.15063 |
| Puducherry | 3.725994 | 0.3 | 0.1 | 0.192101 | 0.194099 | 0.2 | 0.478134 | 0.015057 | 0.2 |
| Punjab | 17.46567 | 0.3 | 0.1 | 0.2 | 0.2 | 0.2 | 0.139514 | 0.007409 | 0.05231 |
| Rajasthan | 43.54541 | 0.3 | 0.1 | 0.2 | 0.199045 | 0.2 | 0.173863 | 0.003455 | 0.012366 |
| Sikkim | 4.184508 | 0.2 | 0.46698 | 0.2 | 0.2 | 0.04 | 0.5 | 0.01308 | 0.2 |
| Tamil Nadu | 1.751252 | 0.283865 | 0.764172 | 0.154537 | 0.040295 | 0.2 | 0.12411 | 0.002323 | 0.156475 |
| Telangana | 50 | 0.3 | 0.117522 | 0.2 | 0.2 | 0.04 | 0.5 | 0.00634 | 0.090637 |
| Tripura | 3.200484 | 0.202165 | 0.899591 | 0.199954 | 0.199863 | 0.069605 | 0.046824 | 0.0016 | 0.06073 |
| Uttar Pradesh | 50 | 0.3 | 0.365293 | 0.040047 | 0.2 | 0.2 | 0.099243 | 0.007514 | 0.01 |
| Uttarakhand | 17.52281 | 0.3 | 0.1 | 0.2 | 0.2 | 0.2 | 0.199483 | 0.009511 | 0.015712 |
| West Bengal | 8.37165 | 0.287528 | 0.551159 | 0.158685 | 0.154366 | 0.2 | 0.133173 | 0.002176 | 0.088322 |

**Supplementary Table S7.**
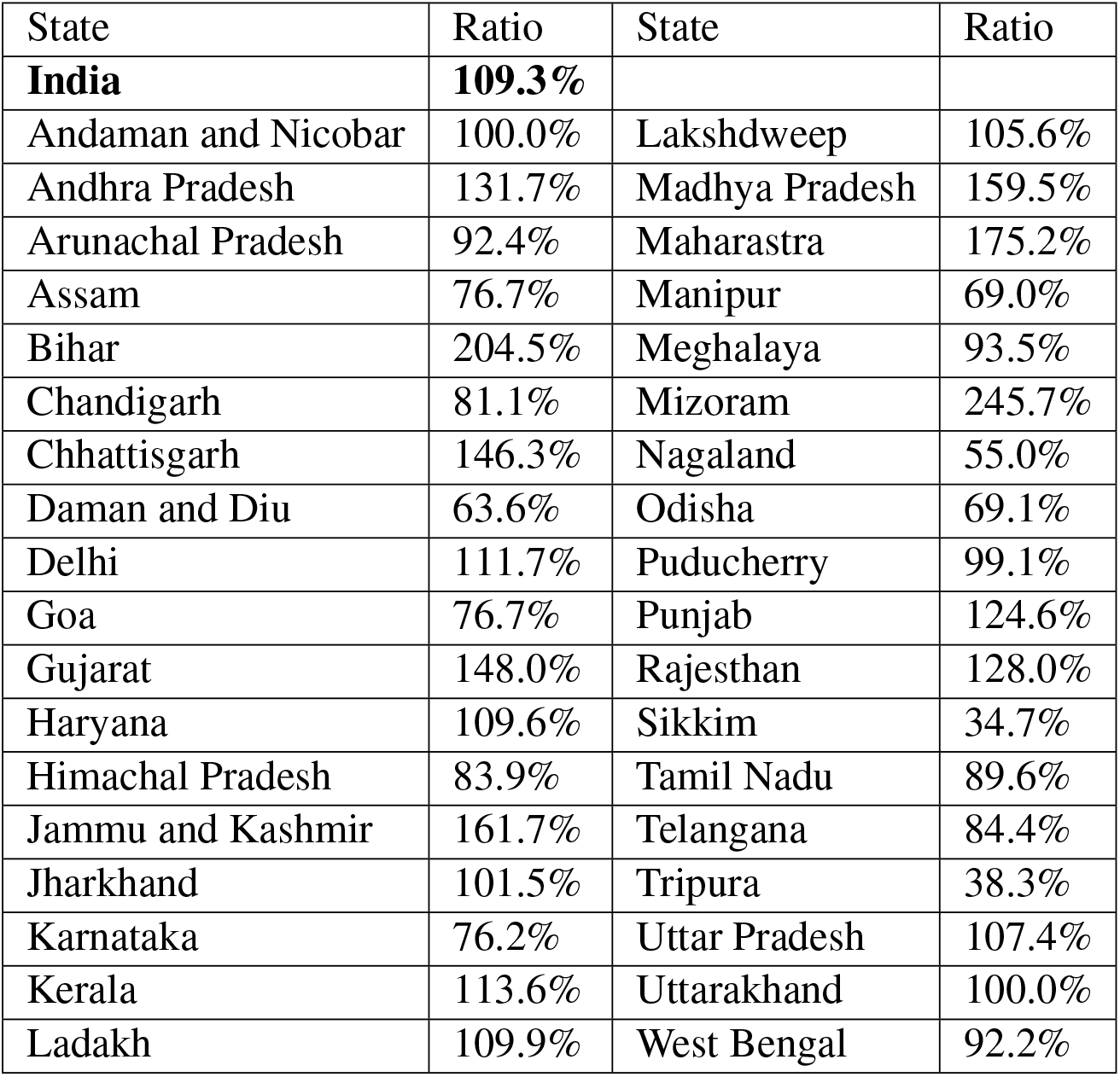
Release ratios for susceptible population: Population release in second lockdown relaxation as a fraction of release in lockdown relaxation in February and March. Release of vaccinated population may be more.

| State | Ratio | State | Ratio |
| --- | --- | --- | --- |
| **India** | **109.3%** |  |  |
| Andaman and Nicobar | 100.0% | Lakshdweep | 105.6% |
| Andhra Pradesh | 131.7% | Madhya Pradesh | 159.5% |
| Arunachal Pradesh | 92.4% | Maharastra | 175.2% |
| Assam | 76.7% | Manipur | 69.0% |
| Bihar | 204.5% | Meghalaya | 93.5% |
| Chandigarh | 81.1% | Mizoram | 245.7% |
| Chhattisgarh | 146.3% | Nagaland | 55.0% |
| Daman and Diu | 63.6% | Odisha | 69.1% |
| Delhi | 111.7% | Puducherry | 99.1% |
| Goa | 76.7% | Punjab | 124.6% |
| Gujarat | 148.0% | Rajasthan | 128.0% |
| Haryana | 109.6% | Sikkim | 34.7% |
| Himachal Pradesh | 83.9% | Tamil Nadu | 89.6% |
| Jammu and Kashmir | 161.7% | Telangana | 84.4% |
| Jharkhand | 101.5% | Tripura | 38.3% |
| Karnataka | 76.2% | Uttar Pradesh | 107.4% |
| Kerala | 113.6% | Uttarakhand | 100.0% |
| Ladakh | 109.9% | West Bengal | 92.2% |

## Notes

### Competing Interest Statement

The authors have declared no competing interest.

### Funding Statement

This study was funded in part by National Science Foundation, USA (Grant No. 2028274)

### Author Declarations

Openly available data from MOHW, India available at: https://github.com/datameet/covid19/blob/master/data/mohfw.json

### Summary of Updates

This version has additional material in section 3 on the impact of lockdowns and methodology in section 4.

